# P2X7-Driven NLRP3 Inflammasome Activation Unveils Novel Serum Biomarkers Associated with the Severity of Mycoplasma pneumoniae Pneumonia in Children

**DOI:** 10.1101/2025.05.09.25327287

**Authors:** Pingping Wang, Wenqian Ding, Lingzhi Wang, Yuhuan Deng, Zixiang Zhan, Shenggang Ding

**Affiliations:** Department of Pediatrics, the First Affiliated Hospital of Anhui Medical University, Hefei, China, 230022; Beijing Children’s Hospital, Capital Medical University, China National Clinical Research Center of Respiratory Diseases, Beijing, China, 100045

**Keywords:** Purinergic receptor P2X7, NOD-like receptor protein 3 (NLRP3), Interleukin-1β (IL-1β), Interleukin-18 (IL-18), Severe mycoplasma pneumoniae pneumonia(sMPP)

## Abstract

**Background:** *Mycoplasma pneumoniae* pneumonia (MPP), often referred to as walking pneumonia, is a common cause of community-acquired pneumonia (CAP) in children and presents with a broad clinical spectrum. While many cases are mild and self-limiting, a subset progresses to severe disease marked by hyperinflammation and systemic complications. This clinical variability presents challenges in early diagnosis and risk stratification. This study aimed to identify prognostic markers associated with MPP severity and explore potential underlying inflammatory mechanisms.

**Methods:** A total of 170 pediatric patients (1–14 years) were enrolled, including 120 MPP cases and 50 non-MPP controls. Serum levels of P2X7, NLRP3, IL-1β, and IL-18 were measured by ELISA. Routine laboratory markers (CRP, LDH, D-dimer, and IgE) were evaluated for their association with severe MPP (sMPP). Pathway enrichment and STRING network analyses were performed to contextualize the clinical markers within inflammation-related molecular pathways. ROC curve analysis was used to assess the predictive value of individual biomarkers.

**Results:** Multivariate logistic regression identified CRP, LDH, D-dimer, and IgE as independent risk factors for sMPP (*p*<0.05). Pathway enrichment revealed these markers to be involved in acute-phase response, coagulation, cytokine signaling, and immune regulation. STRING network analysis further demonstrated their convergence on the NLRP3 inflammasome axis. Serum levels of P2X7, NLRP3, IL-1β, and IL-18 were significantly elevated in sMPP cases compared to non-severe MPP and controls (*p*<0.001). ROC analysis showed all four had strong predictive performance (AUC > 0.7, *p*<0.001).

**Conclusion:** This study confirms that elevated levels of CRP, LDH, D-dimer, and IgE are independently associated with severe *Mycoplasma pneumoniae* pneumonia (sMPP), reflecting systemic inflammation, tissue injury, and immune dysregulation. Importantly, the concurrent upregulation of P2X7, NLRP3, IL-1β, and IL-18 in serum, supported by pathway enrichment and STRING network analyses, highlights a central role for inflammasome activation in sMPP pathogenesis. ROC curve analysis further demonstrated the strong predictive value of these inflammasome-related proteins. Collectively, these findings suggest that P2X7, NLRP3, IL-1β, and IL-18, in combination with conventional markers such as CRP, LDH, D-dimer, and IgE, may serve as valuable biomarkers for the early identification and risk stratification of children at risk for severe MPP, thereby enhancing diagnostic precision and informing clinical decision-making.

## Introduction

In children, Mycoplasma pneumoniae pneumonia (MPP) represents a significant cause of community-acquired pneumonia (CAP), accounting for approximately 10% to 30% of cases [1]. Recent epidemiological trends indicate an increasing prevalence of MPP in younger age groups, along with a rising incidence of severe cases. Although MPP generally has a favorable prognosis, a subset of patients develop severe MPP (sMPP), characterized by persistent high fever, progressive pulmonary deterioration, and multiple pulmonary complications. Additionally, sMPP can manifest with extrapulmonary complications affecting various organ systems, including the gastrointestinal, cardiovascular, hematologic, and urologic systems, further complicating disease management[2]. These clinical challenges highlight the need for early recognition and accurate diagnosis to ensure timely intervention. In routine clinical practice, pediatricians typically rely on laboratory tests, serological antibody detection, and rapid antigen testing to guide MPP management. However, these conventional approaches have limitations, particularly in assessing disease severity with high accuracy. Therefore, the identification of accurate and efficient biomarkers is of substantial clinical significance, as that can enhance diagnostic precision, optimize treatment strategies, and improve prognostic outcomes for children with MPP.

Achieving this goal requires a deeper understanding of the molecular mechanisms underlying the disease’s pathogenesis. However, despite decades of extensive research, the complete pathogenesis of MPP remains incompletely understood. Current evidence suggests that *Mycoplasma pneumoniae* (MP) drives disease progression through a multifaceted interplay of pathogenic mechanisms. These include its ability to adhere to host cells, disrupt cellular homeostasis, and release virulence factors that inflict direct cellular damage. Additionally, MP triggers an exaggerated immune response, leading to the overproduction of inflammatory mediators that contribute to tissue injury. To further complicate its pathogenesis, MP employs sophisticated immune evasion strategies such as adhesion modulation and antigenic mimicry, enabling persistent infection and recurrent disease episodes. These mechanisms highlight the critical role of dysregulated immune responses in the pathogenesis of MPP, particularly in severe cases marked by excessive lung injury, immune hyperactivation, and uncontrolled inflammation.[3].

Notably, a key driver of this process is the activation of the innate immune sensor, NOD-like receptor protein 3 (NLRP3), which serves as a central regulator of host defense. NLRP3 orchestrates the inflammatory pathways by promoting the production and release of pro-inflammatory cytokines such as interleukin-1β (IL-1β) and IL-18. While this response is essential for pathogen clearance, excessive activation can trigger an amplified inflammatory cascade, leading to heightened tissue damage and contributing to disease severity.[4–6]. While the precise molecular pathways underlying NLRP3 inflammasome activation remain incompletely characterized, intracellular potassium (K^+^) efflux is widely recognized as a critical initiating event in most NLRP3 activation pathways [7–9]. Emerging evidence highlights the essential role of the ligand-gated ionotropic purinergic receptor P2X7 in this process, primarily by facilitating rapid K+ efflux in response to pathogenic stimuli [10–12]. Studies suggest that the pathogenesis of severe Mycoplasma pneumoniae pneumonia (sMPP) may be closely linked to excessive activation of the NLRP3 inflammasome, leading to an overproduction of proinflammatory cytokines and subsequent immune hyperactivation [13]. As an upstream activator of NLRP3, P2X7 acts as a key modulator in initiating this inflammatory cascade. However, the potential link between P2X7 and MPP needs to be explored. Additionally, the role of P2X7 receptor modulation in respiratory infections, particularly MPP, remains poorly understood and requires further investigation.

Given the intricate interplay between MP virulence factors, host immune dysregulation, and inflammasome activation, our study assessed circulating levels of inflammasome mediators (P2X7, NLRP3) and inflammatory cytokines (IL-1β, IL-18) in blood serum of pediatric patients to determine their relevance in predicting disease progression and severity, thus evaluating their potential to serve as prognostic markers for MPP disease progression.

## Materials and Methods

### 2.1 Study population

The study enrolled 120 pediatric patients diagnosed with MPP who were admitted to the First Affiliated Hospital of Anhui Medical University between April 2024 to December 2024. Based on clinical diagnostic criteria, the patients were classified into two groups: non-severe MPP (nsMPP, n=65) and severe MPP (sMPP, n=55). The classification of MPP severity was based on the following diagnostic criteria for severe MPP (sMPP) [2]. Patients were classified as sMPP if they exhibited one or more of the following conditions:(I) persistent high fever (>39°C) lasting more than five days or fever persisting for more than seven days without a declining trend in peak temperature;(II) hypoxemia, defined as sustained arterial oxygen saturation (SaO_2_) <92% on room air; (III) increased respiratory and pulse rates, accompanied by clinical signs of respiratory distress and exhaustion, with or without elevated partial pressure of carbon dioxide (PaCO_2_); (IV) radiographic evidence of pulmonary infection, including significant pleural effusion, bronchial casts, vascular occlusion, necrotic lesions, or acute airway obstruction or (V) presence of extrapulmonary complications, such as meningoencephalitis, Guillain-Barré syndrome, myopericarditis, erythema multiforme, or other systemic involvements. To mimimize potential biases, the study adhered to strict inclusion criteria: (I) eligible participants were children aged 1-14 years who met the diagnostic criteria for MPP or sMPP;(II) had negative results from both blood and sputum cultures, and showed no definitive laboratory evidence of viral infection; (III) additionally, informed consent obtained from legal guardians of all the participants. Exclusion criteria included:(I) laboratory-confirmed co-infections;(II) presence of severe underlying chronic conditions such as, cardiopulmonary diseases, hepatic or renal dysfunction, hematologic disorders, immunodeficiency, or inherited metabolic diseases;(III) use of corticosteroids or immunosuppressive agents within four weeks before sample collection;(IV) incomplete medical records;(V) or lack of written informed consent from their legal guardians. Additionally, a control group (CPs) consisting of 50 healthy children who underwent routine health check-ups at the hospital during the same period was included. These children had no history of infections in the preceding month. To ensure comparability, all participants were screened to exclude any immune system disorders and allergic conditions. Venous blood samples were collected from all study participants. The study was conducted following ethical standards and protocols approved by the Ethics Committee of Anhui Medical University.

### 2.2 Measurement of serum proteins

#### 2.2.1 Sample collection

Venous blood samples were collected within 24 hours of admission, before the administration of any treatment. Patients fasted for a minimum of 8 hours before sample collection. Immediately following collection, the blood samples were centrifuged at 3000g for 5 minutes at 4°C to separate plasma components. The resulting supernatant was aliquoted into four portions and stored at −80□ until further analysis.

#### 2.2.2 Experimental methods

Serum levels of P2X7, NLRP3, IL-1β, and IL-18 were quantified using enzyme-linked immunosorbent assay (ELISA). All assays were performed in strict accordance with the manufacturer’s standardized protocols. To ensure technical reproducibility, each measurement was conducted in triplicate. The final reported values represent the mean of three independent experimental replicates.

### 2.3 Pathway Enrichment and Protein–Protein Interaction Analysis

Pathway enrichment analysis was conducted using Metascape (https://metascape.org) to identify biological processes associated with laboratory markers significantly linked to severe MPP. Gene symbols corresponding to CRP, LDHA, FGA, FCER1A, IGHE, and others were analyzed against GO, KEGG, Reactome, and WikiPathways databases. Pathways with *p* < 0.01, enrichment score > 1.5, and ≥3 genes were considered significant. To explore molecular interactions, a protein–protein interaction (PPI) network was constructed using the STRING database (https://string-db.org, version 11.5) with a medium confidence score threshold (≥0.4). Both known and predicted interactions were included. The resulting network revealed connections between inflammation- and immune-related proteins, including central positioning of NLRP3, P2RX7, IL1B, and IL18, guiding subsequent serum protein analyses.

### 2.4 Statistical methods

All statistical analyses were performed using SPSS version 26.0 (IBM Corp., Armonk, NY, USA). GraphPad Prism version 9.5 (GraphPad Software, San Diego, CA, USA) was used for data visualization. Data with normal distribution were presented as mean ± standard deviation (SD). Comparisons between two groups were conducted using independent-samples t-tests, while comparisons among three groups (with homogeneity of variance) were assessed using one-way analysis of variance (ANOVA) followed by Bonferroni post hoc correction. For data not following a normal distribution, results were expressed as median and interquartile range (IQR), and group comparisons were performed using the Mann– Whitney U test (for two groups) or the Kruskal–Wallis H test (for three groups). Categorical variables were reported as frequencies (percentages) and analyzed using the chi-square (χ²) test. Logistic regression was applied to estimate odds ratios (ORs) and corresponding 95% confidence intervals (CIs) for laboratory parameters associated with severe Mycoplasma pneumoniae pneumonia (sMPP). Spearman correlation analysis was used to assess associations between non-normally distributed continuous variables. Receiver operating characteristic (ROC) curve analysis was conducted to evaluate the diagnostic and predictive performance of serum biomarkers for sMPP. A p-value < 0.05 was considered statistically significant.

## Results

### 3.1 Baseline characteristics of study participants across clinical groups

A comparison of gender, age, and weight among children in the control (CP), non-severe Mycoplasma pneumoniae pneumonia (nsMPP), and severe Mycoplasma pneumoniae pneumonia (sMPP) groups revealed no statistically significant differences, as detailed in Table 1. The proportion of male participants was comparable across groups (CP: 54.00%, nsMPP: 53.84%, sMPP: 47.27%). Mean ages were 8.38 ± 1.86, 7.48 ± 2.30, and 7.80 ± 2.42 years, and mean weights were 23.01 ± 6.89, 26.40 ± 9.21, and 27.55 ± 13.07 kg, respectively. These similarities suggest that gender, age, and weight were unlikely to confound subsequent analyses.

**Table 1:** The general comparison of CP, nsMPP, and sMPP.

| Basic Information | CP (n=50) | nsMPP (n=65) | sMPP (n=55) | Std. Error | P value |
| --- | --- | --- | --- | --- | --- |
| Gender (in%) Male | 27 ( 54.00) | 35 ( 53.84 ) | 26 ( 47.27 ) | 0.653 | 0.721 |
| Age (year) | $8.38 \pm 1.86$ | $7.48 \pm 2.30$ | $7.80 \pm 2.42$ | 5.069 | 0.079 |
| Weight (kg) | $23.01 \pm 6.89$ | $26.40 \pm 9.21$ | $27.55 \pm 13.07$ | 3.740 | 0.154 |

### 3.2 Clinical characteristics of study participants across the nsMPP and sMPP groups

A comparative analysis of clinical characteristics revealed several statistically significant differences between the nsMPP and sMPP groups, as shown in Figure 1 and detailed in Supplementary Table 1. Children with sMPP experienced a longer median duration of fever (5 vs. 4 days, *p* < 0.001) and prolonged total disease duration (13 vs. 12 days, *p* = 0.010), along with a longer hospital stay (6 vs. 5 days, *p* = 0.007). Although peak fever temperatures were similar, oxygen saturation was significantly lower in sMPP patients (*p* = 0.002). Pulmonary complications were markedly more common in the sMPP group. Pleural effusion (32.7% vs. 6.2%), lobar pneumonia with consolidation (47.3% vs. 9.2%), and reduced incidence of bronchopneumonia (32.7% vs. 72.3%) were significantly associated with sMPP (all *p* < 0.001). Additionally, the frequency of bronchoscopy treatment (52.7% vs. 7.7%, *p* < 0.001) and glucocorticoid therapy (38.2% vs. 20.0%, *p* = 0.028) was substantially higher in the sMPP group. Extrapulmonary manifestations were also more prevalent in the sMPP group. The proportion of patients without any extrapulmonary complications was significantly lower (63.6% vs. 89.2%, *p* < 0.001), although individual organ involvement (nervous, circulatory, digestive, hematologic, and skin/mucosal) did not differ significantly. These findings underscore the more severe clinical profile of sMPP, characterized by greater pulmonary involvement, extended disease course, and increased need for intervention.

**Fig. 1.**
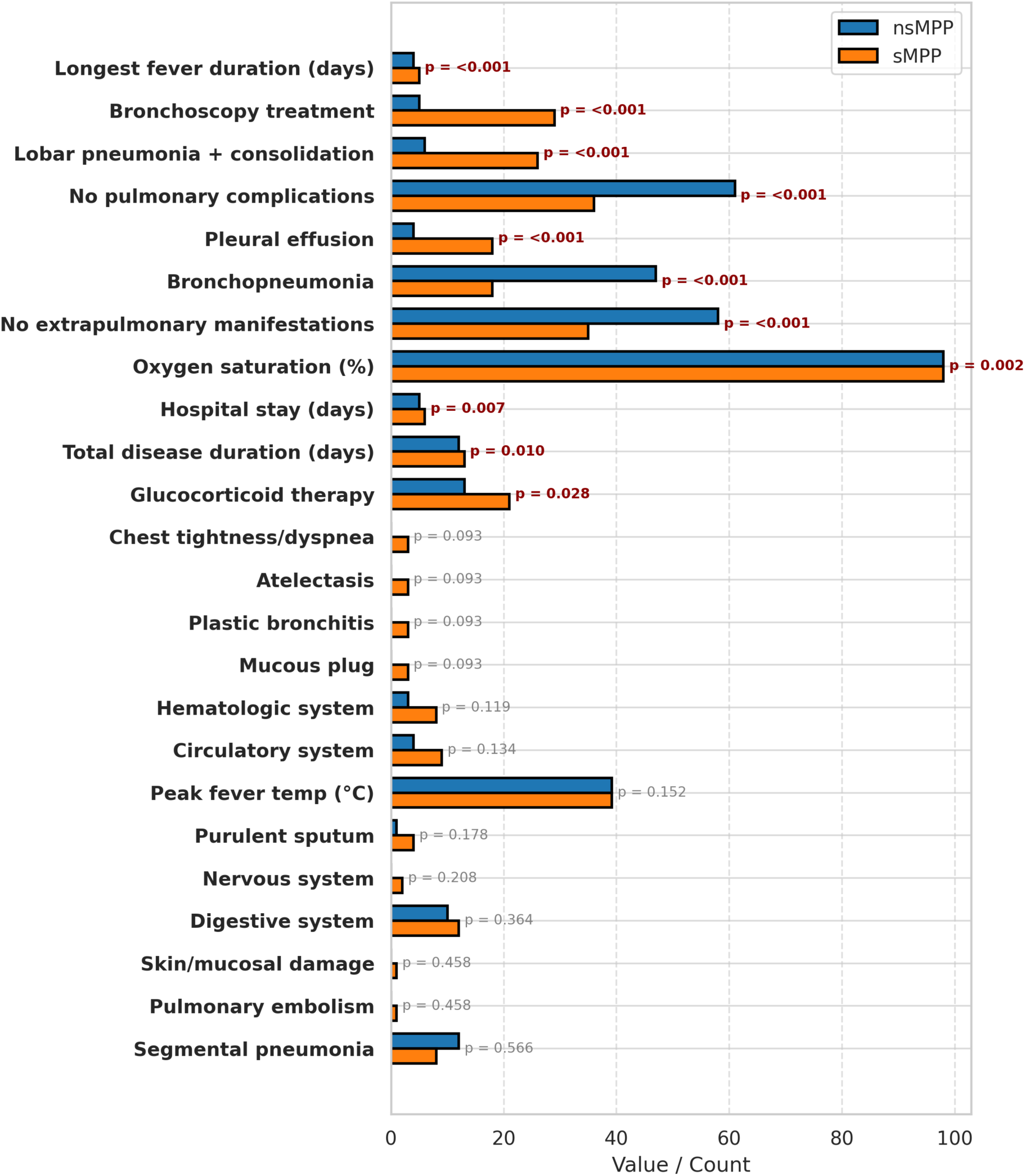
Clinical Differences Between nsMPP and sMPP in Pediatric Patients. Bar chart comparing clinical characteristics of children with non-severe (nsMPP, blue) and severe (sMPP, orange) *Mycoplasma pneumoniae* pneumonia. Values represent counts or averages. Black-outlined bars denote all comparisons; bold red p-values indicate statistically significant differences (p < 0.05).

### 3.3 Serum profiles of routine laboratory indices across nsMPP and sMPP groups

To evaluate differences in routine laboratory parameters, we compared serum indices between children with non-severe (nsMPP) and severe (sMPP) *Mycoplasma pneumoniae* pneumonia. Children in the sMPP group exhibited significantly higher levels of inflammatory and immune markers, including CRP, LDH, D-dimer, and IgE (all *p* < 0.001), reflecting heightened systemic inflammation and immune activation. In contrast, there were no significant differences between groups in WBC count, neutrophil and lymphocyte percentages, eosinophil and basophil ratios, or immunoglobulin levels (IgG, IgA, IgM), as shown in Figure 2A and detailed in Supplementary Table 2 These results suggest that elevated CRP, LDH, D-dimer, and IgE may serve as useful indicators of disease severity in MPP.

**Fig. 2.**
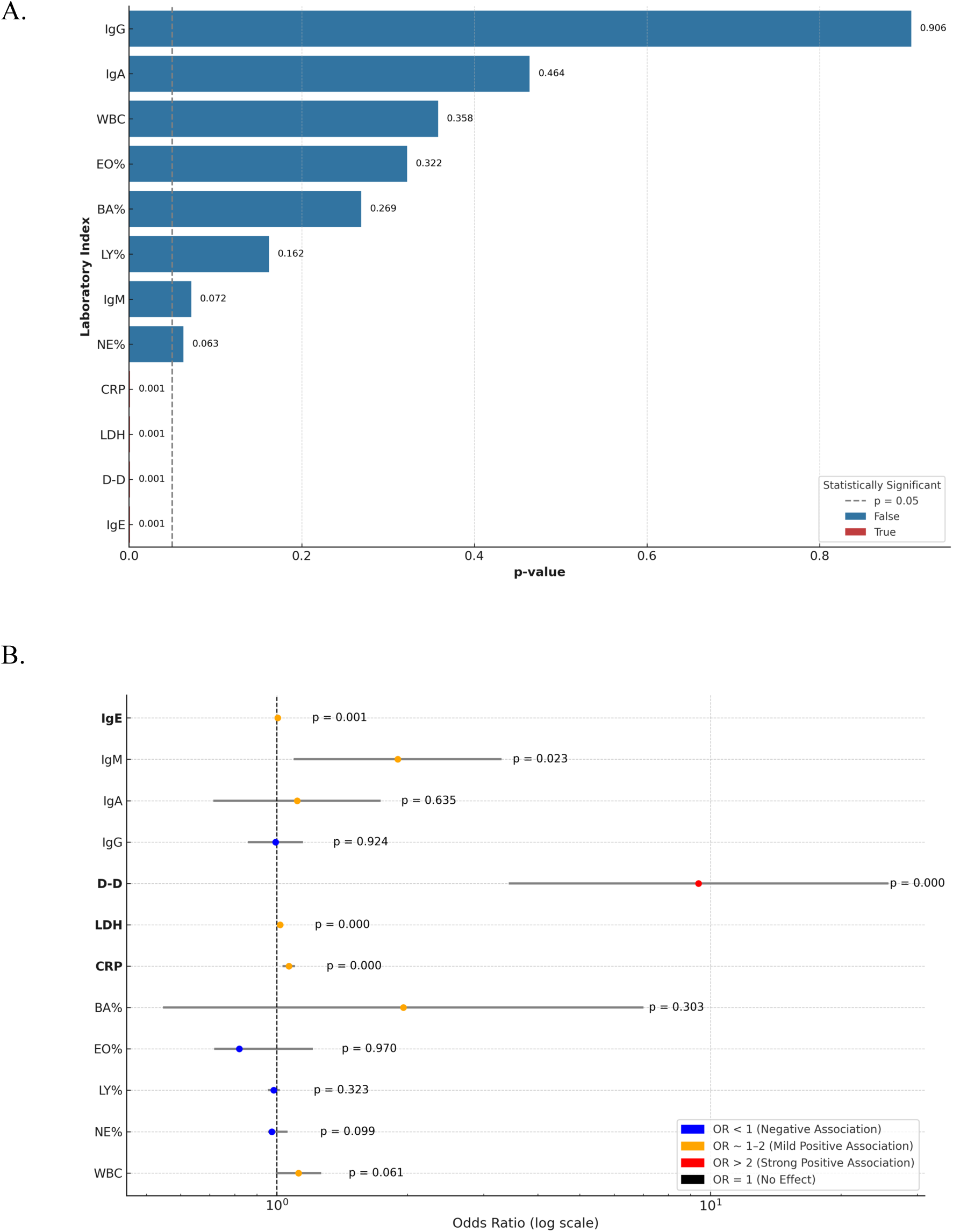
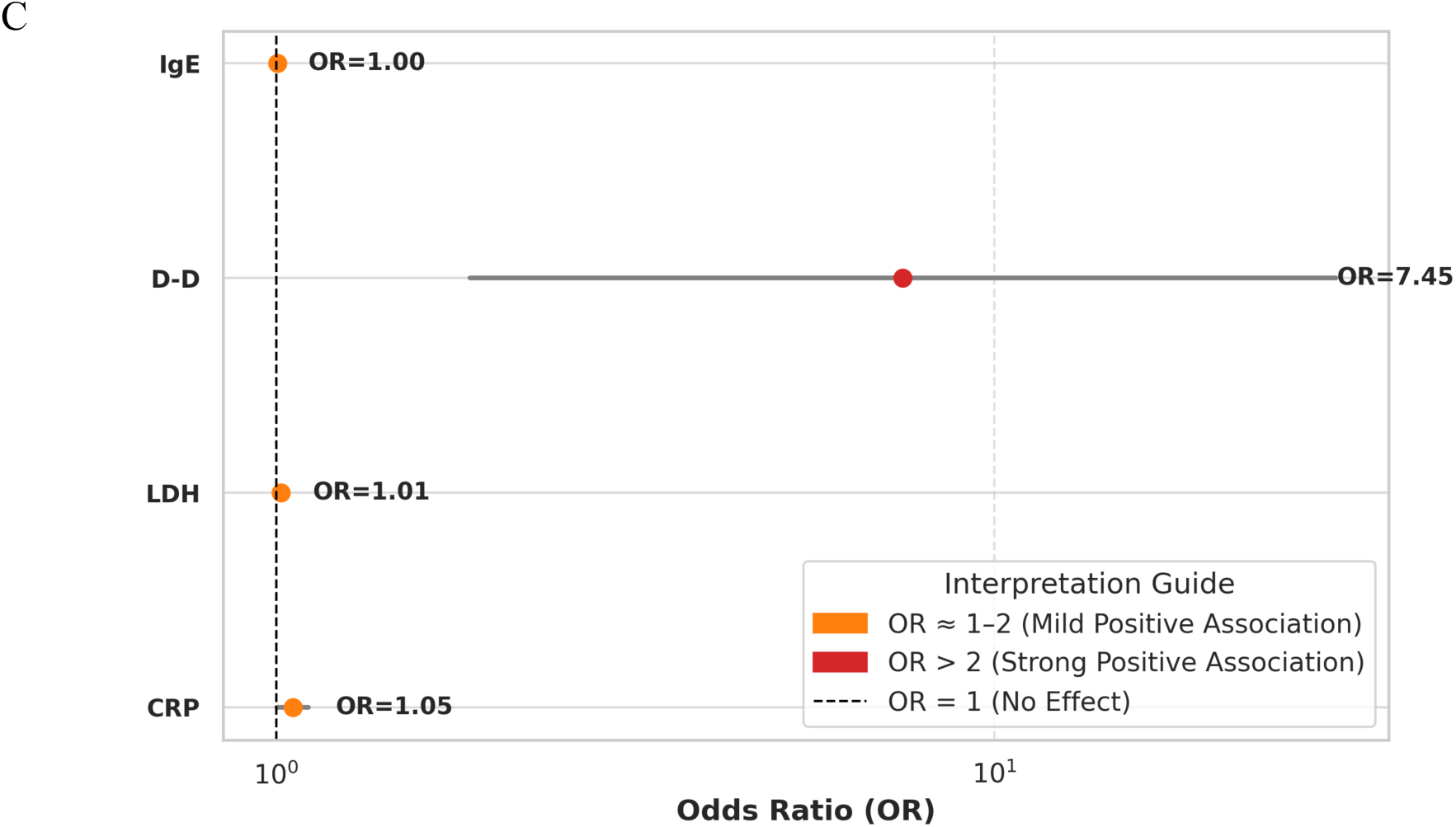
D-dimer exhibits a strong positive association with disease severity in patients, as evidenced by logistic regression and significance testing of laboratory biomarkers. **(A)** Bar plot showing the *p*-values for various laboratory indices in association with disease severity. A dashed vertical line marks the significance threshold (*p* = 0.05). Laboratory parameters with *p* < 0.05 are considered statistically significant. **(B)** Forest plot illustrating odds ratios (OR; log scale) and 95% confidence intervals from univariate logistic regression analysis. Dots represent point estimates and are color-coded: blue (OR < 1, negative association), orange (OR ≈ 1–2, mild positive association), and red (OR > 2, strong positive association). **(C)** Focused forest plot of the top four significant laboratory predictors (IgE, CRP, LDH, and D-dimer), with D-dimer showing the strongest association (OR = 7.45). All ORs were computed using logistic regression models. Statistical significance was set at *p* < 0.05.

### 3.3 Inflammatory, Coagulation, and Immune Markers Predict Severe MPP in Univariate Analysis

Univariate binary logistic regression analysis was performed to evaluate the association between routine laboratory parameters and the risk of severe *Mycoplasma pneumoniae* pneumonia (sMPP). Elevated levels of CRP, LDH, D-dimer, and IgE were significantly associated with sMPP (*p* < 0.05), as shown in Figure 2B and detailed in supplementary Table 3. In contrast, other parameters, including WBC count, neutrophil and lymphocyte percentages, eosinophil and basophil percentages, IgG, and IgA, were not significantly associated with disease severity. These findings highlight the potential of inflammatory, coagulation, and immune markers as predictors of severe MPP.

### 3.4 Multivariate Analysis Identifies Inflammatory and Coagulation Markers as Independent Predictors of sMPP

Laboratory parameters that were identified as statistically significant in the univariate analysis were subsequently included in a multivariate logistic regression analysis to determine their independent association with sMPP development. The multivariate analysis confirmed that the elevated levels of CRP, LDH, D-dimer (D-D), and IgE remained independently and significantly (*p*<0.05) associated with the development of severe MPP, as shown in Figure 2C and supplementary Table 4. These findings suggest that elevated levels of inflammatory and coagulation markers, along with increased IgE, serve as important independent risk factors for disease severity.

### 3.5. Pathway Enrichment Analysis Reveals Inflammatory Signatures Linked to Inflammasome Activation in sMPP

Pathway enrichment analysis was conducted using the laboratory markers significantly associated with sMPP to further interpret the clinical relevance of these findings. As shown in Figure 3A, these biomarkers were enriched in key immune and inflammatory pathways, including the acute-phase response, complement and coagulation cascades, cytokine signaling, and immunoglobulin-mediated immune responses. Notably, CRP, D-dimer, and IgE contributed prominently to systemic inflammation and complement activation pathways, highlighting their involvement in the pathogenesis of severe MPP. These pathway-level insights provide a mechanistic basis for examining inflammasome-related proteins, which are central mediators of innate immune activation and pyroptotic inflammation. The alignment between clinical biomarkers and enriched inflammatory pathways supports the hypothesis that inflammasome activation plays a key role in sMPP and justifies its targeted investigation in subsequent analyses.

**Fig. 3.**
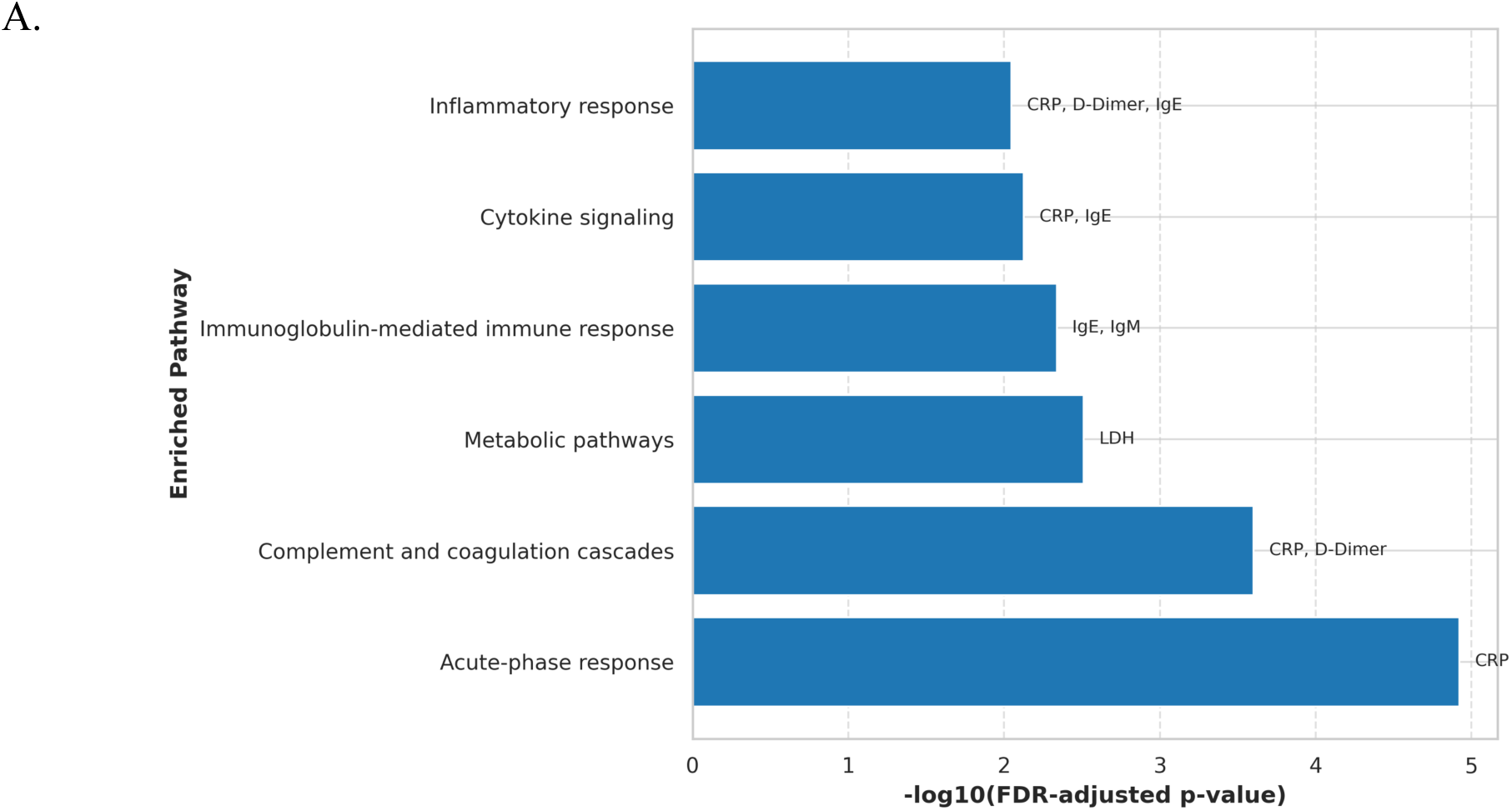
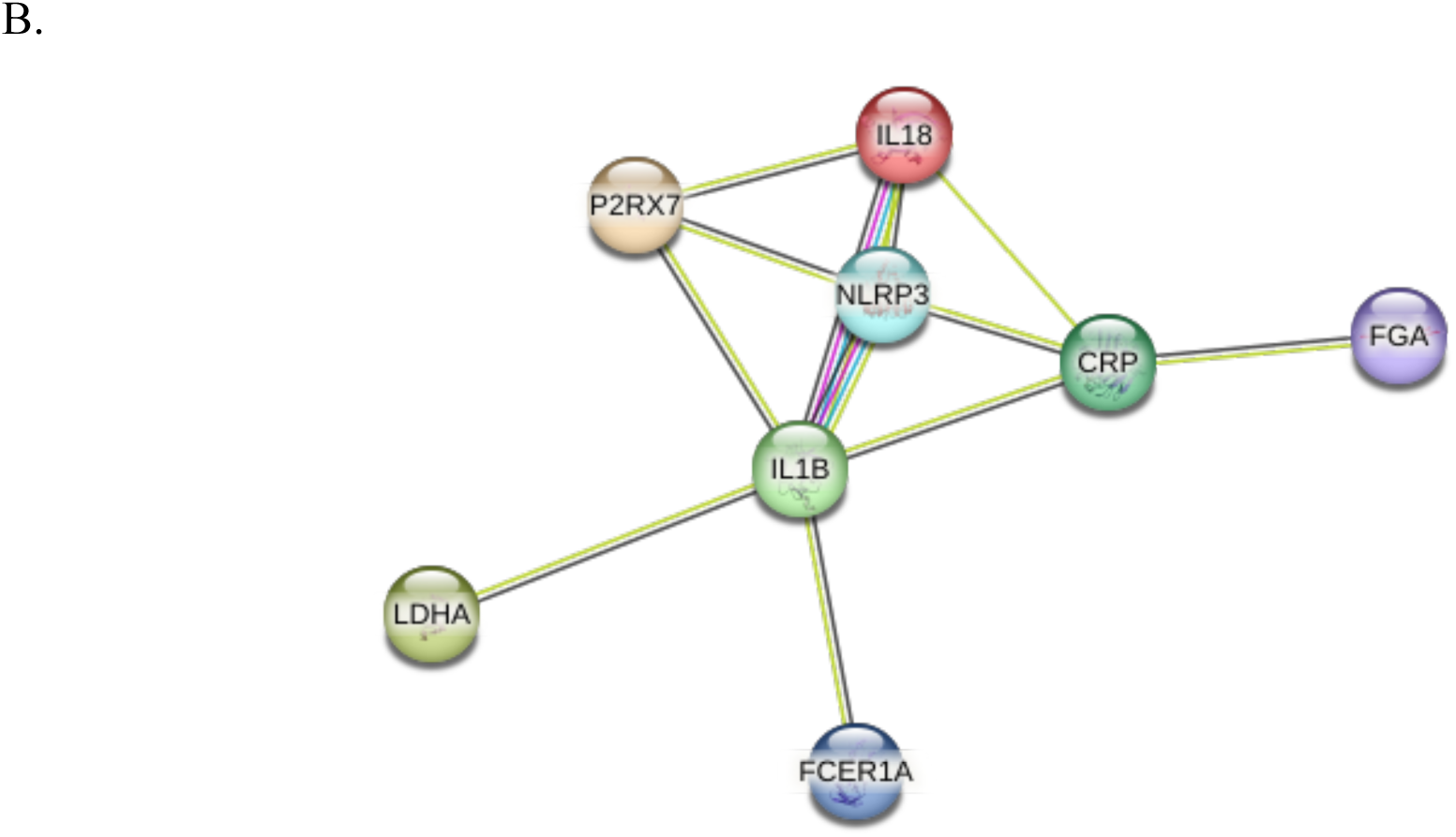
Acute-phase and inflammatory pathways enriched in severe *Mycoplasma pneumoniae* pneumonia (sMPP) are driven by CRP-associated responses and P2RX7-NLRP3 inflammasome signaling. **(A)** Bar plot showing enriched pathways associated with severity-linked biomarkers. The x-axis represents pathway enrichment as –log[[(FDR-adjusted *p*-value), with higher values indicating stronger significance (*FDR* < 0.05). Biomarkers contributing to each pathway are labeled. **(B)** Protein–protein interaction network illustrating functional links among severity-associated proteins, including CRP, LDHA, FCER1A, and inflammatory mediators (NLRP3, IL1B, IL18). Edge colors represent evidence types, and thickness indicates interaction confidence. Results highlight the P2RX7–NLRP3– IL1B axis as a potential driver connecting innate immune activation with acute-phase and immunoglobulin-mediated responses in sMPP.

### 3.6 STRING Network Analysis Supports the Role of Inflammasome in sMPP Pathogenesis

To further investigate the molecular context of the laboratory markers associated with severe MPP, a protein-protein interaction network was generated using the STRING database, as shown in Figure 3B. This network revealed functional associations between CRP, LDHA, FGA (related to D-dimer), and FCER1A (related to IgE), all of which converged on the NLRP3 inflammasome axis. NLRP3 was centrally connected to its upstream activator P2RX7 and downstream effectors IL-1β and IL-18, indicating a coherent inflammatory signaling module. CRP was linked to both NLRP3 and IL-1β, while LDHA was connected to IL-1β, reflecting roles in inflammasome priming and pyroptotic cell death, respectively. FCER1A showed a direct interaction with IL-1β, suggesting a potential link between IgE-mediated responses and inflammasome activity. These connections provide a mechanistic framework linking the elevated clinical markers observed in sMPP with the NLRP3 inflammasome pathway. Based on this network and the pathway enrichment results, we next assessed serum levels of P2X7, NLRP3, IL-1β, and IL-18 to evaluate inflammasome activation in the context of disease severity.

### 3.7 Serum analysis confirms that Activation of the P2X7-NLRP3 Inflammasome Pathway Correlates with Disease Progression

Given the importance of these inflammatory markers in MPP progression, we next investigated and measured the serum levels of P2X7, NLRP3 in CPs, nsMPPs, and sMPPs. The Kruskal–Wallis H-test was employed to assess differences among the three groups. Results showed that both P2X7 and NLRP3 levels were significantly elevated in nsMPPs and sMPPs compared to CPs (p < 0.001), with sMPPs exhibiting significantly higher levels than nsMPPs, as shown in Figure 4. These data suggest a robust activation of the P2X7-NLRP3 pathway in sMPP compared to nsMPP and CP. Based on these findings, we next evaluated the serum concentrations of the downstream inflammatory cytokines IL-1β and IL-18 using one-way ANOVA and F-tests. A similar trend was observed: both IL-1β and IL-18 levels increased progressively from CPs to nsMPPs and were highest in sMPPs, highlighting a consistent trend of escalating inflammatory response as the disease becomes more severe.

**Fig. 4.**
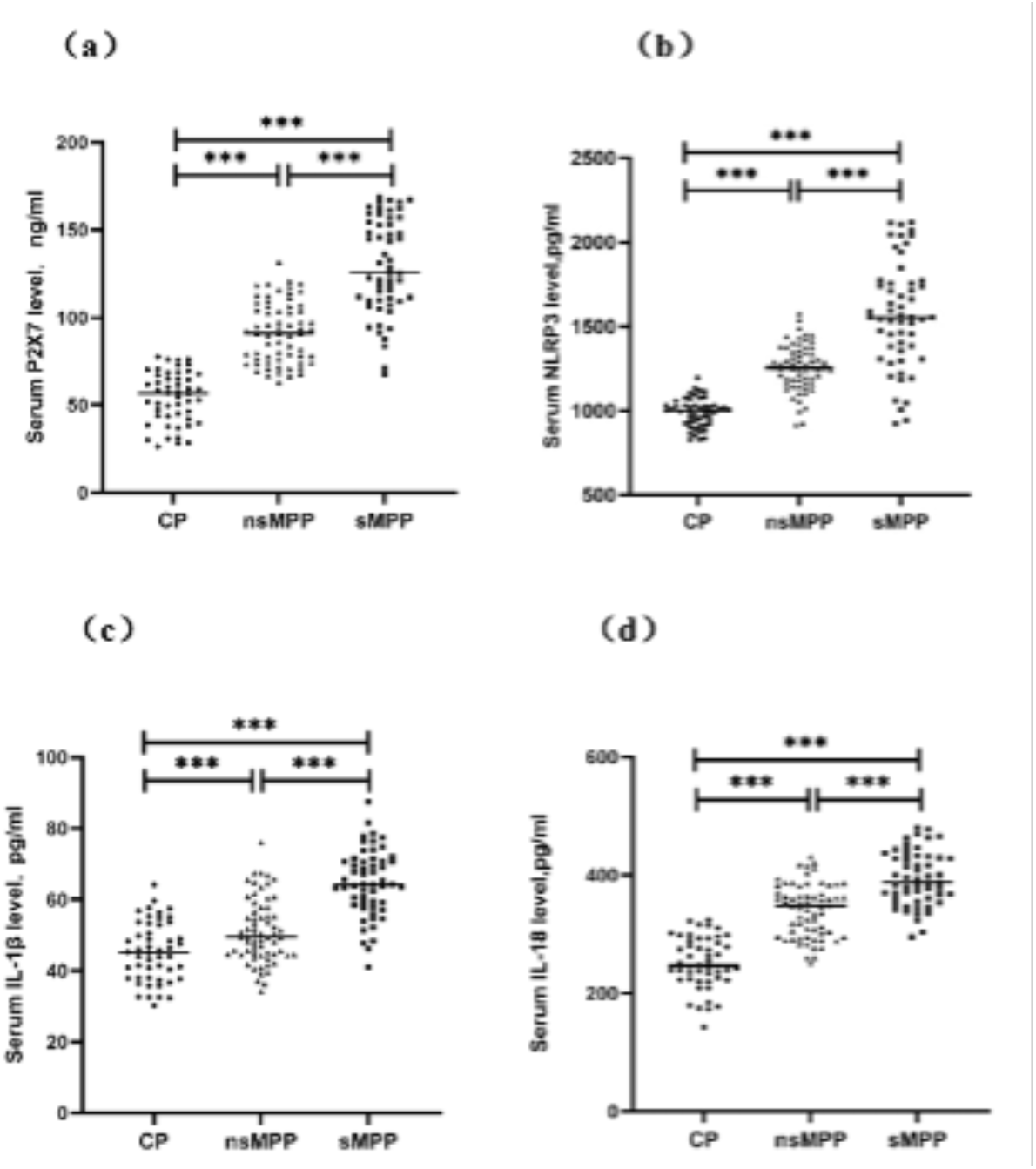
Serum levels of P2RX7, NLRP3, IL-1β, and IL-18 are significantly elevated in children with severe *Mycoplasma pneumoniae* pneumonia (sMPP). Serum concentrations of (a) P2RX7, (b) NLRP3, (c) IL-1β, and (d) IL-18 were measured in children with community-acquired pneumonia (CP), non-severe MPP (nsMPP), and severe MPP (sMPP). All cytokines showed progressive increases from CP to sMPP. Data are presented as scatter plots with means ± SD. Comparisons between groups were performed using one-way ANOVA followed by Tukey’s post hoc test. \*\**p* < 0.001 was considered statistically significant.

### 3.8 Positive Correlations Among Inflammasome Components Highlight a Coherent Inflammatory Signature

Building on the observed progressive increases in serum P2X7, NLRP3, IL-1β, and IL-18 levels across the groups, we further examined the interrelationships among these biomarkers. We conducted a Spearman correlation analysis to investigate the associations between serum P2X7, NLRP3, IL-1β, and IL-18, as well as between NLRP3 and the cytokines IL-1β and IL-18. The analysis revealed significant positive correlations: serum P2X7 levels were strongly correlated with NLRP3 (r = 0.8071), IL-1β (r = 0.6587), and IL-18 (r = 0.7593), as shown in Fig.5 A–C. Additionally, serum NLRP3 levels showed robust positive correlations with both IL-1β (r = 0.7371) and IL-18 (r = 0.8391), as shown in Fig.5 D-E. All correlations were statistically significant (p < 0.001), underscoring the tight interplay within the P2X7-NLRP3 inflammasome pathway and its downstream effectors in the context of MPP severity.

**Fig. 5.**
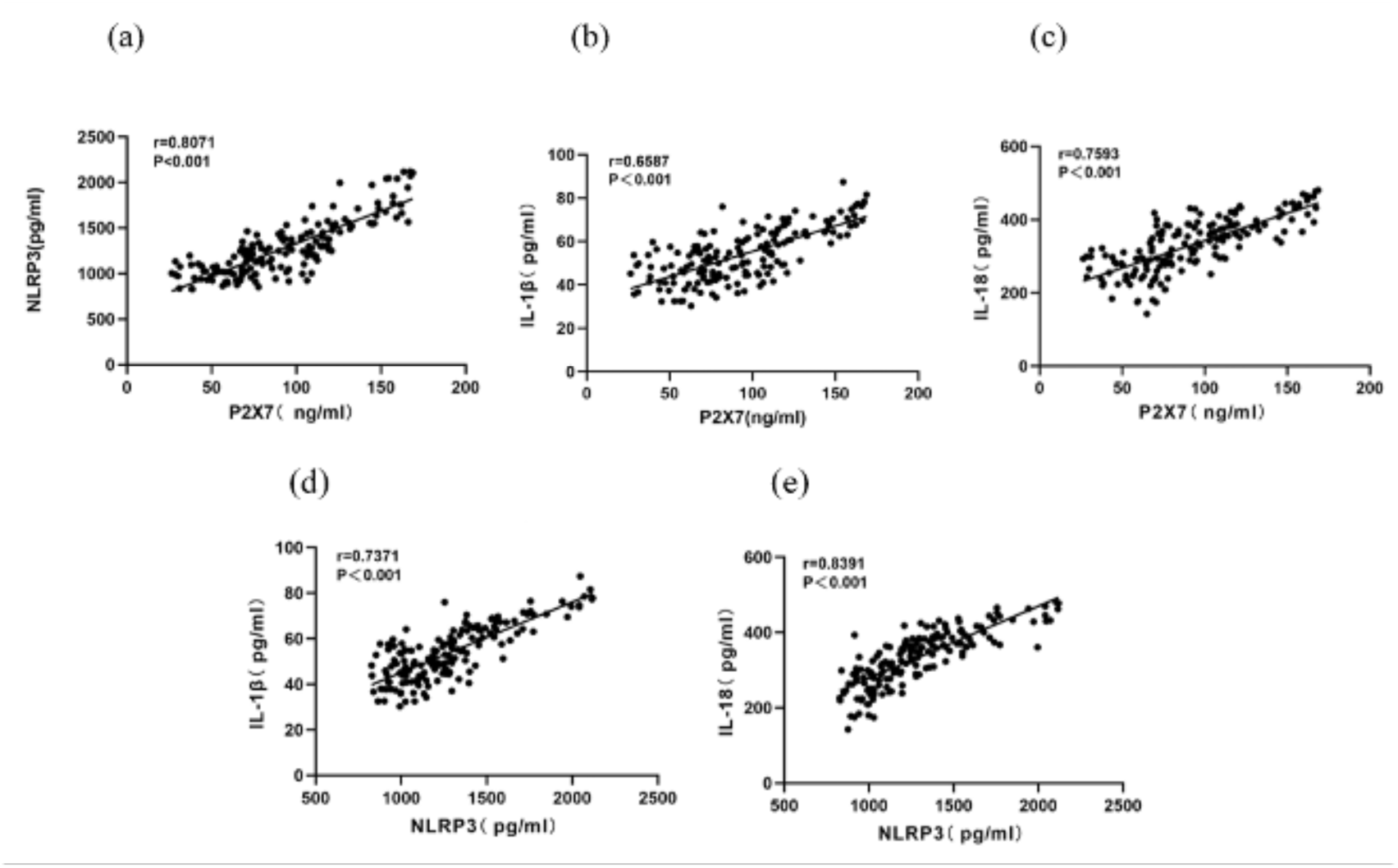
Serum P2RX7 levels strongly correlate with NLRP3, IL-1β, and IL-18 expression in children with *Mycoplasma pneumoniae* pneumonia (MPP). Pearson correlation analysis demonstrating positive associations between serum P2RX7 levels and (a) NLRP3, (b) IL-1β, and (c) IL-18 concentrations. Similarly, NLRP3 levels were positively correlated with (d) IL-1β and (e) IL-18 levels. Each dot represents an individual patient sample. Correlation coefficients (*r*) and *p*-values are indicated in each panel. Statistical significance was defined as *p* < 0.05.

### 3.9 Serum P2X7, NLRP3, IL-1**β**, and IL-18 can be used as Reliable Biomarkers for Identifying Severe MPP in Children

To assess the prognostic utility of serum P2X7, NLRP3, IL-1β, and IL-18 in differentiating between non-severe (nsMPP) and severe Mycoplasma pneumoniae pneumonia (sMPP) in children, Receiver Operating Characteristic (ROC) analysis was performed. This statistical method is widely employed to evaluate the diagnostic or predictive performance of biomarkers by plotting sensitivity (true positive rate) against 1-specificity (false positive rate) across a range of threshold values. A central metric derived from ROC analysis is the Area Under the Curve (AUC), which reflects the overall discriminative capacity of the test. An AUC of 1.0 denotes perfect discrimination, whereas an AUC of 0.5 indicates no better accuracy than random chance. Our results showed that all four biomarkers demonstrated AUC values exceeding 0.7, signifying good predictive performance (p < 0.001). These results suggest that elevated serum levels of P2X7, NLRP3, IL-1β, and IL-18 are potential prognostic biomarkers for identifying children at increased risk of developing severe MPP, as shown in Fig. 6 and detailed in Table 2.

**Fig. 6.**
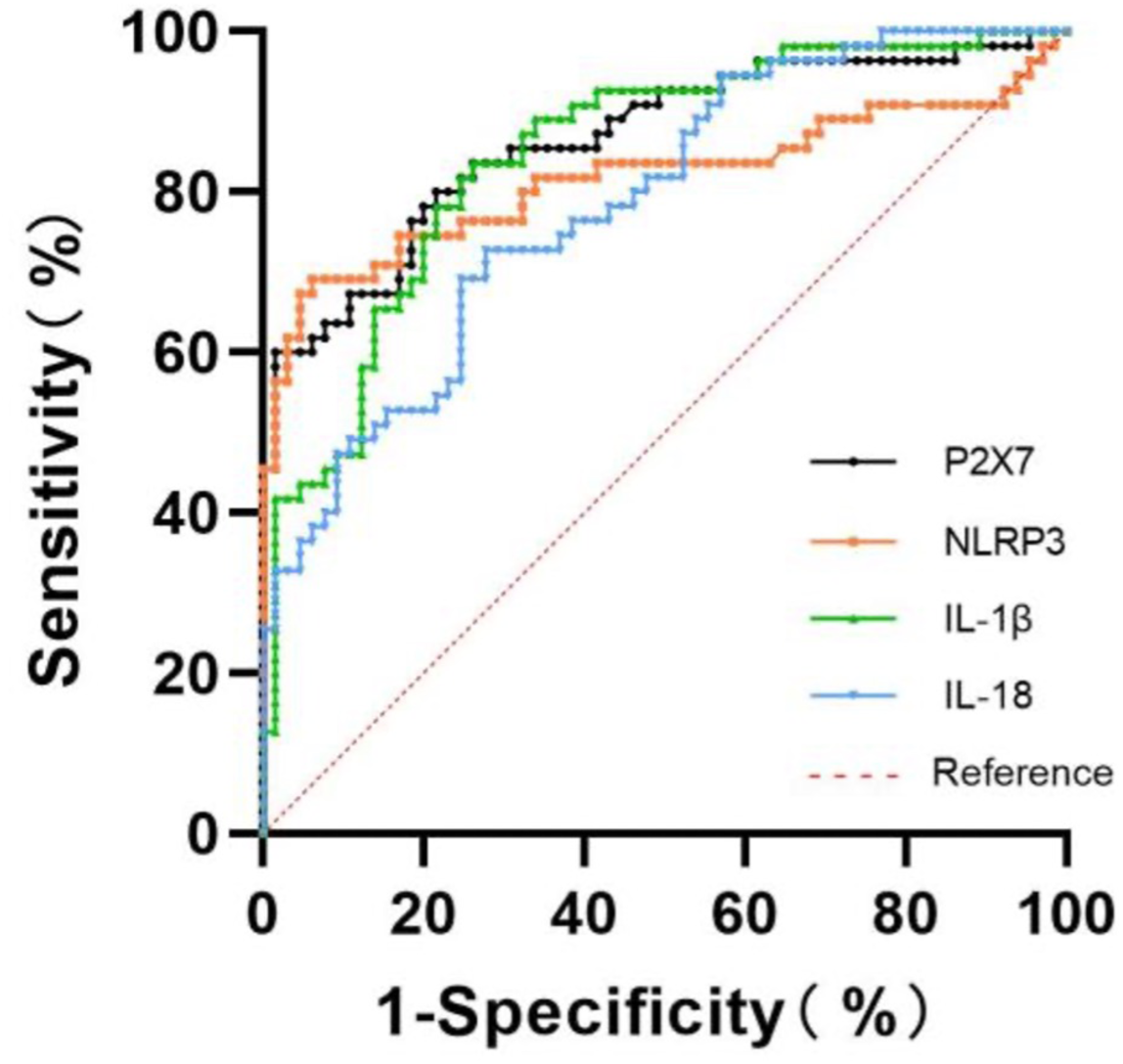
P2RX7, NLRP3, IL-1β, and IL-18 show strong predictive value for severe *Mycoplasma pneumoniae* pneumonia (sMPP). ROC curves assessing the ability of serum P2RX7, NLRP3, IL-1β, and IL-18 to distinguish sMPP from non-severe cases. The y-axis indicates sensitivity, and the x-axis shows 1–specificity. The red dashed line represents the reference (AUC = 0.5). All markers displayed high AUCs, reflecting strong diagnostic performance.

**Table 2.** ROC Curve Analysis of P2X7, NLRP3, IL-1β, and IL-18 for predicting sMPP.

| Indicators | AUC | 95% CI | Cut-off point | Youden index | Sensitivity (%) | Specificity (%) | *p* value |
| --- | --- | --- | --- | --- | --- | --- | --- |
| P2X7 | 0.866 | 0.791~0.921 | 106.980 (ng/ml) | 0.585 | 80.00 | 78.46 | <0.001 |
| NLRP3 | 0.817 | 0.737~0.882 | 1448.985 (pg/ml) | 0.629 | 69.09 | 93.85 | <0.001 |
| IL-1β | 0.847 | 0.770~0.906 | 55.333 (pg/ml ) | 0.575 | 83.64 | 73.85 | <0.001 |
| IL-18 | 0.785 | 0.701~0.855 | 365.165 (pg/m) | 0.450 | 72.73 | 72.31 | <0.001 |

## Discussion

Mycoplasma pneumoniae (MP) is a common respiratory pathogen known to primarily infect the mucosal epithelium of both the upper and lower airways, often resulting in considerable tissue damage. MP infections can progress to Mycoplasma pneumoniae pneumonia (MPP), a widespread condition that affects individuals across all age groups but is particularly common in children, with the highest incidence typically observed in late summer and early autumn. This heightened susceptibility in pediatric populations is influenced by several factors, including the immaturity of the immune system, anatomical differences in the respiratory tract, and increased exposure risks in communal environments such as schools and daycare centers [18].

Despite its prevalence, early diagnosis of MP infection remains challenging due to its non-specific clinical manifestations, which include fever, cough, and sore throat, symptoms that overlap with other respiratory infections. Laboratory diagnostic methods, such as nucleic acid testing and serological antibody assays, are commonly employed; however, their sensitivity in the early stages of infection is limited by low MP load and insufficient antibody levels. Additionally, radiological findings in the early phase of MPP often lack distinct characteristics, further complicating differential diagnosis. These diagnostic limitations frequently result in delayed intervention, increasing the risk of disease progression to severe MPP (sMPP). Given these constraints, the identification of more reliable, early-stage biomarkers for sMPP remains a critical challenge.

In this study, we confirmed that elevated levels of inflammatory, coagulation, and immune markers, specifically C-reactive protein (CRP), lactate dehydrogenase (LDH), D-dimer (D-D), and immunoglobulin E (IgE), were significantly associated with disease severity in pediatric MPP. These biomarkers reflect key pathological processes in severe MPP, including systemic inflammation, cellular injury, and endothelial dysfunction. Our findings are consistent with previous reports that support the use of these markers in routine clinical practice for evaluating disease progression. Despite their widespread clinical use, each of these markers has inherent limitations. CRP, while indicative of systemic inflammation, lacks disease specificity and is subject to individual variability, potentially leading to mismatches between CRP levels and actual disease severity [14]. LDH, a marker of cellular damage, may not show significant elevation during the early stages of infection, limiting its predictive utility [15]. Similarly, D-D levels increase upon activation of coagulation and fibrinolysis but demonstrate a delayed rise, reducing their effectiveness for early severity assessment [16]. IgE, although linked to immune responses, does not directly correlate with pulmonary lesion severity or respiratory impairment and shows irregular fluctuations throughout disease progression [17]. Moreover, the absence of a standardized protocol for the combined use of these markers further complicates objective severity assessment [19].

However, when we further tried to elucidate the underlying mechanisms of MPP pathogenesis, we realized the role of immune activation and inflammatory mediators in disease progression. It is well established that *Mycoplasma pneumoniae* initiates infection by triggering host immune responses, leading to the release of inflammatory cytokines. In severe cases, this response becomes excessive, exacerbating tissue damage and contributing to increased disease severity [20–22]. Supporting these observations, our pathway enrichment and protein-protein interaction analyses revealed the involvement of inflammasome multiprotein complexes, particularly the NLRP3 inflammasome, as central regulators of the inflammatory cascade. The NLRP3 inflammasome is one of the most well-characterized innate immune complexes, comprising the sensor protein NLRP3, the adaptor ASC (apoptosis-associated speck-like protein containing a CARD), and the effector caspase-1 [17]. To investigate the relevance of inflammasome activation in sMPP, we quantified serum levels of P2X7, NLRP3, IL-1β, and IL-18 in pediatric patients with non-severe MPP (nsMPP) and severe MPP (sMPP), comparing them to healthy controls (CP). Our results revealed a progressive elevation of these inflammatory markers from the CP group to the nsMPP group and further to the sMPP group. These results represent distinct stages of NLRP3 inflammasome activation: P2X7 as a trigger of potassium efflux, NLRP3 as the inflammasome sensor, and IL-1β and IL-18 as downstream inflammatory cytokines. Normally, the NLRP3 inflammasome remains inactive in immune cells. However, upon encountering pathogen-associated molecular patterns (PAMPs) or damage-associated molecular patterns (DAMPs), it undergoes conformational changes, leading to its activation. This process triggers the secretion of IL-1β and IL-18, amplifying the inflammatory cascade and contributing to pyroptotic cell death [3–5]. The P2X7 receptor, a trimeric ionotropic receptor belonging to the purinergic P2 family, is widely expressed in innate and adaptive immune cells. Studies have demonstrated that P2X7 activation induces intracellular K+ efflux, a key stimulus for NLRP3 inflammasome assembly [23–24]. This leads to the activation of caspase-1 and the subsequent maturation of IL-1β and IL-18. IL-1β activates the nuclear factor-kappa B (NF-κB) signaling cascade, promoting the transcription of pro-inflammatory genes, whereas IL-18 stimulates interferon-γ production, driving a type I inflammatory response via T-helper 1 cells [25–27]. These cytokines serve as critical regulators of inflammation, and their excessive production is implicated in the progression of MPP to severe disease. Previous studies on P2X7 receptor involvement in respiratory inflammation have primarily focused on chronic airway diseases such as chronic obstructive pulmonary disease and asthma. However, its role in MP-induced inflammatory responses remains largely unexplored. Notably, there are currently no clinical studies linking P2X7 and the NLRP3/IL-1β/IL-18 axis with MPP, highlighting a significant gap in research. Further, our Spearman correlation analysis demonstrated a strong positive correlation between P2X7 levels and serum concentrations of NLRP3, IL-1β, and IL-18, confirming the upstream regulatory role of P2X7 in NLRP3 inflammasome activation. Additionally, serum NLRP3 levels were significantly correlated with both IL-1β and IL-18, reinforcing the mechanistic link between inflammasome activation and disease severity.

Furthermore, ROC curve analysis demonstrated that each of the inflammasome-related proteins, P2X7, NLRP3, IL-1β, and IL-18, exhibited strong predictive value for identifying children at risk of developing severe MPP. Among these, P2X7 showed the highest area under the curve (AUC), followed by IL-1β, NLRP3, and IL-18. Notably, while NLRP3 had slightly lower sensitivity, it exhibited high specificity, making it particularly valuable for confirming severe disease. Collectively, these results suggest that inflammasome activation is not only mechanistically involved in the pathogenesis of sMPP but may also serve as a clinically useful axis for disease stratification and targeted intervention.

Despite the promising implications of our findings, several limitations should be acknowledged. First, although we excluded patients with recent infections or pneumonia caused by known pathogens other than *Mycoplasma pneumoniae*, the possibility of undetected co-infections cannot be entirely ruled out and may have influenced the results. Second, our analysis was limited to serum biomarker levels, which may not fully capture localized inflammatory responses within lung tissue. Future studies incorporating tissue-specific analyses could offer deeper insights and help identify additional diagnostic targets for severe MPP. Finally, as all participants in this study were Asian children from Southeast China, the generalizability of our findings to broader, more diverse populations may be limited. Multi-center studies with larger, ethnically diverse cohorts are warranted to validate and extend these observations.

In summary, this study demonstrates that children with Mycoplasma pneumoniae pneumonia (MPP) have significantly higher serum levels of P2X7, NLRP3, IL-1β, and IL-18 compared to healthy controls, underscoring the role of P2X7-mediated inflammasome activation in the pathogenesis of MPP. We found a strong correlation between the serum concentrations of these markers and disease severity, indicating that severe MPP (sMPP) is associated with an intensified inflammatory response. Additionally, these markers emerged as potential predictors of sMPP. Taken together, our findings suggest that serum levels of P2X7, NLRP3, IL-1β, and IL-18 may serve as useful biomarkers for assessing MPP severity in pediatric patients.

## Data Availability

All data produced in the present work are contained in the manuscript

## Statements and Declarations

## Abbreviations

WBC: White blood cell count
NE%: Neutrophil percentage
LY%: Lymphocyte percentage
EO%: Eosinophil percentage
BA%: Basophil percentage
a: refers to z-score
b: corresponds to the t-value

## Competing interests

The authors declare no competing interests related to the content of this article. The authors have no relevant financial or non-financial interests to disclose.

## Author contributions

S.D. had full access to all study data and takes responsibility for the integrity and accuracy of the data analysis, including any adverse effects. P.W., W.D., L.W., Y.D., and Z.Z. made substantial contributions to the study design, data analysis and interpretation, and manuscript preparation.

## Funding

This work was supported by the General Program of the National Natural Science Foundation of China (Grant No. 52273113).

## Ethics approval

This study was performed in line with the principles of the Declaration of Helsinki. Approval was granted by the Clinical Research Ethics Committee of The First Affiliated Hospital of Anhui Medical University **(29.03.2024 /Approval No Quick-PJ2024-04-50)**.

## Consent to participate

As this was a retrospective study, informed consent from parents or legal guardians was waived with approval from the Medical Research Ethics Committee of Anhui Medical University. Human Ethics and Consent to Participate declarations: not applicable.

## Notes

### Competing Interest Statement

The authors have declared no competing interest.

### Funding Statement

This study was funded by the General Program of the National Natural Science Foundation of China (Grant No. 52273113)

### Author Declarations

Ethics committee of The First Affiliated Hospital of Anhui Medical University gave ethical approval for this work

