## Supplementary Tables for "P2X7-Driven NLRP3 Inflammasome Activation Unveils Novel Serum Biomarkers Associated with the Severity of Mycoplasma pneumoniae Pneumonia in Children"

**Supplementary Table 1. Clinical characteristics of study participants**

| Clinical Characteristics | nsMPP | sMPP | P-value |
|---|---|---|---|
| Longest duration of fever (days) | 4 | 5 | <0.001 |
| Peak fever temperature (°C) | 39.20 | 39.20 | 0.152 |
| Chest tightness/dyspnea, n (%) | 0 | 3 | 0.093 |
| Oxygen saturation (%) | 98.00 | 98.00 | 0.002 |
| None (pulmonary complications), n (%) | 61 | 36 | <0.001 |
| Pleural effusion, n (%) | 4 | 18 | <0.001 |
| Pulmonary embolism, n (%) | 0 | 1 | 0.458 |
| None (extrapulmonary manifestations), n (%) | 58 | 35 | <0.001 |
| Nervous system, n (%) | 0 | 2 | 0.208 |
| Circulatory system, n (%) | 4 | 9 | 0.134 |
| Digestive system, n (%) | 10 | 12 | 0.364 |
| Hematologic system, n (%) | 3 | 8 | 0.119 |
| Skin and mucosal damage, n (%) | 0 | 1 | 0.458 |
| Bronchopneumonia, n (%) | 47 | 18 | <0.001 |
| Segmental pneumonia, n (%) | 12 | 8 | 0.566 |
| Lobar pneumonia + consolidation, n (%) | 6 | 26 | <0.001 |
| Atelectasis, n (%) | 0 | 3 | 0.093 |
| Glucocorticoid therapy, n (%) | 13 | 21 | 0.028 |
| Bronchoscopy treatment, n (%) | 5 | 29 | <0.001 |
| Plastic bronchitis (PB), n (%) | 0 | 3 | 0.093 |
| Purulent sputum, n (%) | 1 | 4 | 0.178 |
| Mucous plug, n (%) | 0 | 3 | 0.093 |
| Length of hospital stay (days) | 5 | 6 | 0.007 |
| Total disease duration (days) | 12 | 13 | 0.010 |

**Supplementary Table 2 Comparison of laboratory indices between nsMPP and sMPP groups**

| **Laboratory indices** | **nsMPP** | **sMPP** | ***P value*** |
|---|---|---|---|
| WBC（$\times 10^9$/L） | 7.32(5.52,8.72) | 7.30(5.86,11.93) | 0.358 |
| NE% (%) | 57.96±10.62 | 61.69±13.67 | 0.063 |
| LY% (%) | 32.27±10.44 | 30.15±12.94 | 0.162 |
| EO% (%) | 1.50(0.60,2.90) | 1.20(0.20,3.24) | 0.322 |
| BA% (%) | 0.30(0.20,0.40) | 0.30(0.20,0.50) | 0.269 |
| CRP/(mg/L) | 10.35(3.65,15.74) | 24.55(9.87,41.17) | ＜0.001 |
| LDH（U/L） | 320(269.25,374.5 | 334(283.75,432.50) | ＜0.001 |
| D-D (ug/ml) | 0.74(0.57,1.07) | 1.19(0.75,2.20) | ＜0.001 |
| IgG(g/L) | 9.27(7.88,10.58) | 9.27(7.59,10.50) | 0.906 |
| IgA(g/L) | 1.30(0.75,1.92) | 1.37(0.91,2.07) | 0.464 |
| IgM(g/L) | 1.15(0.79,1.48) | 1.27(0.93,1.83) | 0.072 |
| IgE(g/L) | 87.50(27.50,199.7 | 127.00(58.00,502.50 | ＜0.001 |

**Supplementary Table 3 Univariate Binary Logistic Regression Analysis**

| **Laboratory indices** | **Standard Error** | **Wald** | **OR** | **95% CI** | ***p* value** |
|---|---|---|---|---|---|
| WBC（$\times 10^9$/L） | 0.061 | 3.507 | 1.122 | 0.995~1.265 | 0.061 |
| NE% ( %) | 0.016 | 2.727 | 0.974 | 0.995~1.059 | 0.099 |
| LY% ( %) | 0.016 | 0.975 | 0.984 | 0.953~1.016 | 0.323 |
| EO% ( %) | 0.100 | 0.001 | 0.819 | 0.717~1.212 | 0.970 |
| BA% ( %) | 0.651 | 1.062 | 1.957 | 0.546~7.014 | 0.303 |
| CRP/(mg/L) | 0.017 | 14.323 | 1.066 | 1.031~1.102 | 0.000 |
| LDH（U/L） | 0.004 | 21.240 | 1.017 | 1.010~1.024 | 0.000 |
| D-D (ug/ml) | 0.514 | 18.964 | 9.393 | 3.427~25.740 | 0.000 |
| IgG(g/L) | 0.075 | 0.009 | 0.993 | 0.857~1.150 | 0.924 |
| IgA(g/L) | 0.227 | 0.225 | 1.114 | 0.714~1.737 | 0.635 |
| IgM(g/L) | 0.282 | 5.189 | 1.900 | 1.094~3.301 | 0.023 |
| IgE(g/L) | 0.001 | 10.929 | 1.004 | 1.002~1.006 | 0.001 |

**Supplementary Table 4**: Multivariate Binary Logistic Regression Analysis

| **Laboratory indices** | **Standard Error** | **Wald** | **OR** | **95% CI** | ***p* value** |
|---|---|---|---|---|---|
| CRP/(mg/L) | 0.026 | 4.274 | 1.055 | 1.003~1.110 | 0.039 |
| LDH（U/L） | 0.005 | 8.544 | 1.015 | 1.005~1.025 | 0.003 |
| D-D (ug/ml) | 0.708 | 8.046 | 7.451 | 1.860~29.843 | 0.005 |
| IgE(g/L) | 0.002 | 6.448 | 1.005 | 1.001~1.009 | 0.011 |
